# Epidemiology of *Vibrio Cholerae* Infections in the Households of Cholera Patients in the Democratic Republic of the Congo: PICHA7 Prospective Cohort Study

**DOI:** 10.1101/2024.12.16.24318937

**Authors:** Christine Marie George, Presence Sanvura, Alves Namunesha, Jean-Claude Bisimwa, Kelly Endres, Willy Felicien, Camille Williams, Shubhanshi Trivedi, Kilee L. Davis, Jamie Perin, David A. Sack, Justin Bengehya, Ghislain Maheshe, Cirhuza Cikomola, Lucien Bisimwa, Daniel T. Leung, Alain Mwishingo

**Author notes:** **Corresponding Author:** Christine Marie George, PhD, Professor, Department of International Health, Program in Global Disease Epidemiology and Control, Johns Hopkins Bloomberg School of Public Health, 615 N. Wolfe Street, Room E5535, Baltimore, Maryland 21205-2103. Co-first author.

## Abstract

**Background:** The aim of this prospective cohort study is to build evidence on transmission dynamics and risk factors for *Vibrio cholerae* infections in cholera patient households.

**Methods:** Household contacts of cholera patients were observed for 1-month after the index cholera patient was admitted to a health facility for stool, serum, and water collection in urban Bukavu in South Kivu, Democratic Republic of the Congo. A *V. cholerae* infection was defined as a *V. cholerae* bacterial culture positive result during the 1-month surveillance period and/or a four-fold rise in a *V. cholerae* O1 serological antibody from baseline to the 1-month follow-up.

**Results:** Twenty-seven percent of contacts (134 of 491) of cholera patients had a *V. cholerae* infection during the surveillance period. Twelve percent (9 of 77) of cholera patient households had a stored water sample with *V. cholerae* by bacterial culture, and 7% (5 of 70) had a water source sample with *V. cholerae*. Significant risk factors for symptomatic *V. cholerae* infections among contacts were stored food left uncovered (Odds Ratio (OR): 2.39, 95% Confidence Interval (CI): 1.13, 5.05) and younger age (children <5 years) (OR: 2.09, 95% CI: 1.12, 3.90), and a drinking water source with >1 colony forming unit *E*.*coli* / 100mL (OR: 3.59, 95% CI: 1.46, 8.84) for *V. cholerae* infections.

**Conclusions:** The findings indicate a high risk of cholera among contacts of cholera patients in this urban cholera endemic setting, and the need for targeted water treatment and hygiene interventions to prevent household transmission of *V. cholerae*.

**Summary:** In this prospective cohort study in the Democratic Republic of the Congo, the majority of cholera patient households had multiple *Vibrio cholerae* infected household members and both source water and stored drinking water samples had *V. cholerae*.

## Introduction

Worldwide there are estimated to be 2.9 million cholera cases annually.^1^ In 2024, there were major cholera outbreaks in 14 African countries._2_ Climate change has increased droughts and floods in Sub-Saharan Africa through events such as El Niño which have increased cholera outbreaks to historic highs in this region.^2,3^ The Democratic Republic of the Congo (DRC) has one of the highest rates of cholera in Africa._2_ Risk factors for *Vibrio cholerae* infections in previous studies include having an unimproved water source, storing drinking water without a cover, unimproved sanitation, and lack of hygiene practices.^4,5^ These studies indicate suboptimal water, sanitation, and hygiene (WASH) are important cholera transmission routes.

Individuals living in close proximity to cholera patients are at an increased risk of subsequent *V. cholerae* infections.^6-8^ Previous studies in rural and urban Bangladesh have found that household contacts of cholera patients had a 100 times higher risk of cholera then the general population during the 7-day period after the cholera patient is admitted at a health facility for treatment.^8,9^ Risk factors for *V. cholerae* infections among the household contact of cholera patients include having *V. cholerae* in the household’s water source or stored drinking water, consuming street vended food, O blood group status, and younger age.^8-10^ All previous published studies of household transmission of *V. cholerae* to date are from Bangladesh and India. There are no household contact studies of cholera in sub-Saharan Africa despite the high rates of cholera in this region, highlighting the need for evidence on household transmission of *V. cholerae* in sub-Saharan Africa.^2^

The aim of this prospective cohort study is to build evidence on transmission dynamics of *V. cholerae* in cholera patient households, and to understand WASH risk factors for *V. cholerae* infections that can be targeted in future interventions. This evidence will inform the delivery of cholera control strategies in the DRC to ensure interventions are targeting risk factors for *V. cholerae* infections for those residing in high-risk transmission hotspots for cholera around cholera patients.

## Methods

### Ethical approval

This study was conducted in urban Bukavu in the South Kivu province of the DRC. We received ethical approval for this study from the institutional review boards of the Johns Hopkins School of Public Health and Catholic University of Bukavu. Written informed consent was obtained from all participants or their guardians.

### Study Design

This prospective cohort study of household contacts of cholera patients was conducted from December 2021 to December 2023. Screening of diarrhea patients for *V. cholerae* was conducted daily at 115 healthcare facilities in Bukavu, DRC. Diarrhea patients were recruited if the following criteria was met: 1) admitted to a health facility with three or more loose stools over a 24-hour period; 2) had no running water inside of their home (mostly informal settlements); and 3) planned to reside in Bukavu for the next month. Diarrhea patients were tested for cholera using the Crystal VC O1 rapid diagnostic test with results confirmed by bacterial culture for *V. cholerae*.^11^ Cholera patients were defined as diarrhea patients with a stool bacterial culture result positive for *V. cholerae*. Household members of the cholera patient were eligible for the cohort study if: 1) they shared the same cooking pot and resided in the same home with the cholera patient for the last three days; and 2) planned to reside with the cholera patient for the next month. Eligible household contacts were enrolled within 24 hours of patient enrollment. The sample size for the number of cholera patient households was determined by the number of cholera patients that could be screened and were willing to participate in the cohort study from December 2021 to December 2023.

Cholera patient households were visited 1, 3, 5, 7, 9, and 11 days and 1-month after the index cholera patient in the household was admitted to a health facility to conduct clinical surveillance and spot checks. Whole stool (all timepoints) and blood samples (baseline and 1-month follow-up) were also collected for *V. cholerae* and *Escherichia coli* bacterial culture and serological analyses. During clinical surveillance visits, a questionnaire was administered to obtain demographic information on diarrhea (3 or more loose stools over a 24-hour period), age, and gender. An unannounced spot check (to prevent households preparing for our arrival) was conducted at each timepoint to: (1) collect a sample of the household’s water source and stored drinking water to test for free chlorine and *V. cholerae* and *E. coli*; (2) check for the presence of soap in the household (a proxy measure of handwashing with soap behavior^12^); and (3) check the covering status of stored drinking water and stored food in the home. The World Health Organization (WHO) free chlorine cutoff of >0.2 mg/L for safe drinking water relative to chlorine was used^13^⍰ Chlorine was measured using a digital colorimeter (Hach, Loveland, CO, USA). The WHO guideline for drinking water quality of <1 colony forming units (CFU) /100 mL of *E*.*coli* in drinking water was used as the cutoff to define safe drinking water quality relative to microbial contamination^14^⍰ Information was also collected at baseline on household water source and sanitation option type using the categories defined by the WHO/ UNICEF Joint Monitoring Program^15,16^⍰

### Laboratory Analyses

All water, stool and blood samples were brought to the PICHA7 Enteric Disease Microbiology Laboratory within three hours of when the sample was produced (stool) or collected (serum) for *V. cholerae* and *E. coli* bacterial culture and serological analyses. One hundred milliliters of household water source and stored water was analyzed for *E*.*coli* by bacterial culture using standard microbiological methods published elsewhere.^17^ For *V. cholerae* analyses, four hundred milliliters of water samples were filtered through a 0.22 µm nitrocellulose membrane filter. The filter was then transferred to a vial containing three ml of APW broth and incubated at 37° C for 18 hours. Likewise, four hundred μl of the watery portion from each patient’s stool or 2-3 grams of whole stool was transferred to a vial using a scoop (no swab was used) containing three ml of alkaline peptone water (APW) broth and kept at 37°C for 6-18 hours. After enrichment, 5–10 μl of APW broth from both water and stool was streaked onto Thiosulphate Citrate Bile Sucrose agar then incubated at 37° C for 18–24 hours. Presumptive colonies were sub-cultured on gelatin agar and incubated at 37° C for 18–24 hours.^18^ *V. cholerae* colonies from gelatin agar plates were tested to determine their serogroups using slide agglutination with polyvalent antiserum, followed by serogroup O1 specific antisera testing as previously published.^19^ Blood samples were analyzed for blood group type using the agglutination method.^20^ Serum was separated from blood and frozen at -80°C. Serum samples were shipped to the United States and analyzed for IgG and IgA antibodies to *V. cholerae* O1 Ogawa and Inaba O-specific polysaccharide (OSP), and IgG to cholera toxin B subunit (CTB) using enzyme-linked immunosorbent assay (ELISA) adapted from previously published methods (see Supplemental File 1 for additional detail).^21^

### Statistical Analysis

The study primary outcomes were: (1) a household contact with a *V. cholerae* infection defined as a positive bacterial culture result during the surveillance period and/or a 4-fold rise in serum *V. cholerae* O1 OSP IgG, IgA, or CTB IgG antibody (a serological marker for infection) from baseline to the 1-month follow-up, and (2) a household contact with a symptomatic *V. cholerae* infection, defined as a *V. cholerae* infection (using the definition described above) with diarrhea during the 1-month surveillance period. Univariate logistic regression models were performed using generalized estimating equations (GEE) to account for clustering within households and estimate the odds of developing a *V. cholerae* infection. The predictors were household and individual level risk factors summarized over the surveillance period. O blood group status and doxycycline comparisons were performed using chi square and fisher exact tests. All analyses were performed using SAS, version 9.4 (SAS Institute Inc., Cary, NC, USA).

## Results

From December 2021 to December 2023, 87 cholera patients with 491 household contacts were followed prospectively. These 87 cholera patients were recruited from 16 health facilities during epidemiological surveillance. There were 83 index cholera patient households, 4 households had >1 index cholera patient. Fifty-five percent of index cholera patients (48/87) and 56% (275/491) of household contacts were female (Table 1). The median age of index cholera patients was 14 years and 13 years for contacts of cholera patients. The median number of individuals in cholera patient households was 8 ± 3 (standard deviation) (range: 3-16).

**Table 1.** Individual Level Characteristics of Cholera Patient Households.

|  | All Participants |  | Index Patients |  | Household Contacts<sup>1</sup> |  |
| --- | --- | --- | --- | --- | --- | --- |
|  | % | n | % | n | % | n |
| **Participants** | 578 |  | 87 |  | 491 |  |
| **Baseline Age** |  |  |  |  |  |  |
| Median ± SD (Min - Max) (Years) | 13 ± 16 (0-83) | | 14 ± 18 (0-79) | | 13 ± 15 (0-83) | |
| 0-5 Years | 19% | 112 | 16% | 14 | 20% | 98 |
| 5-14 Years | 34% | 194 | 33% | 29 | 34% | 165 |
| 14 Years or Greater | 47% | 272 | 51% | 44 | 46% | 228 |
| **Female Gender** | 56% | 323 | 55% | 48 | 56% | 275 |
| **Blood Group Status** |  |  |  |  |  |  |
| O Blood Group | 45% | 222 | 51% | 37 | 44% | 185 |
| A Blood Group | 31% | 152 | 28% | 20 | 31% | 132 |
| B Blood Group | 21% | 103 | 15% | 11 | 22% | 92 |
| AB Blood Group | 3% | 17 | 6% | 4 | 3% | 13 |
| **Reported Baseline Antibiotic Usage (past 48 hours)** | 13% | 71 | 34% | 30 | 9% | 41 |
| **Reported Antibiotic Usage During the 1-Month Surveillance Period** | 42% | 245 | 84% | 73 | 35% | 172 |
| **Reported Baseline Oral Rehydration Solution (ORS) Usage (past 48 hours)** | 17% | 89 | 88% | 75 | 3% | 14 |
| **Reported ORS Usage During the 1-Month Surveillance Period** | 21% | 122 | 93% | 81 | 8% | 41 |
| **Baseline Intravenous Fluids** | 13% | 69 | 69% | 59 | 2% | 10 |
| **Intravenous Fluids During the 1-Month Surveillance Period** | 15% | 85 | 78% | 68 | 4% | 17 |

1. Household contacts were defined as individuals sharing the same cooking pot and residing in the same home with the cholera patient for the last three days and who also planned to reside with the cholera patient for the next month.

Thirty-four percent of cholera patients (30/87) reported consuming antibiotics within 48 hours prior to enrollment with 3% (3/87) consuming doxycycline (the standard of care for cholera in DRC). Fifty-one percent of index cholera patients (37/72) had an O blood group status compared to 44% of household contacts of patients (185/422) (p<0.0001). Ninety-six percent (473/491) of household contacts provided a blood sample and 99% (489/491) provided a stool sample during the 1-month surveillance period. Individual level characteristics of household contacts stratified by whether they had a bacterial culture or serological result available is reported in Supplementary Table 1. Ninety-three percent of households (77/83) were present during an unannounced spot check visit during the surveillance period.

Sixty-seven percent of cholera patient households (56/83) had >1 *V. cholerae* infected household contacts during the 1-month surveillance period (Table 2). Thirty-seven percent of households (31/83) had >1 contact with a symptomatic *V. cholerae* infection. Forty-two percent of households (35/83) had >1 *V. cholerae* infected contact. Twelve percent of households had a stored water sample (9/77) with *V. cholerae* and 7% had a source water sample (5/70) with *V. cholerae*. Sixteen percent of households (12/77) had a positive water sample for *V. cholerae* (stored or source) during the surveillance period.

**Table 2.**
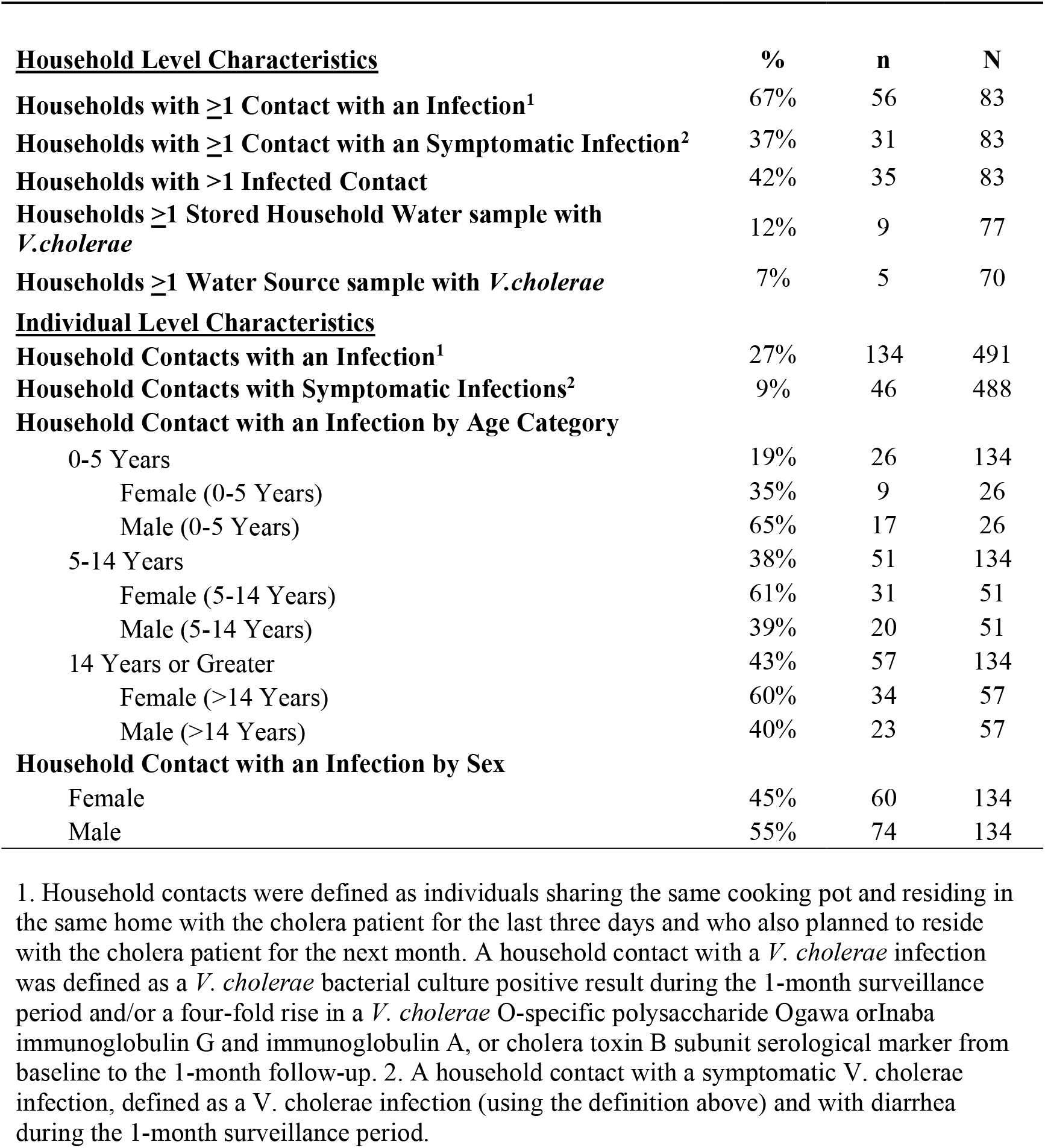
Household and Individual level *V. cholerae* Infection and Water Sample Characteristics for Household Contacts of Cholera Patients.

For individual level *V. cholerae* infection characteristics in cholera patient households, 27% (134/491) of household contacts had a *V. cholerae* infection during the 1-month surveillance period. Nine percent of contacts (46/488) had a symptomatic *V. cholerae* infection. Nineteen percent (26/134) of *V. cholerae* infections were among individuals <5 years, 38% (51/134) for 5-14 years, and 43% (56/134) for >14 years. Five percent of contacts (6/134) with a *V. cholerae* infection reported visiting a health facility for the treatment of diarrhea. Of all household contacts positive for V. cholerae, 70% were positive by bacterial culture (93/134), 41% were positive by serology (55/134), and 10% were positive by both (14/134). For bacterial culture-defined infections, 38% of *V. cholerae* infections were first detected on Day 1 (35/93), 27% (25/93) on Day 3, 19% (17/93) on Day 5, 6% (6/93) on Day 7, 9% (8/93) on Day 9, 1% on Day 11 (1/93), and 1% (1/93) at Month-1. The median duration of shedding of *V. cholerae* for contacts with an initial *V. cholerae* infection after Day 1 was 2 days ± 1.76 (standard deviation) (range: 1-7).

Twenty-six percent (112/435) of household contacts had stored food in their household uncovered at all spot check visits. Thirty-one percent of household contacts (148/478) resided in households with stored drinking water with <0.2 mg/L free chlorine at all spot check visits, and 47% (200/422) for source water. Sixty-two percent of household contacts (287/463) resided in households with basic water service and 27% (126/463) for basic sanitation service. Twenty-three percent (14/60) of contacts resided in a household with *E*.*coli* in a drinking water source, and 46% (31/68) with *E*.*coli* in stored drinking water during the surveillance period. Significant risk factors for symptomatic *V. cholerae* infections among contacts were stored food left uncovered (Odds Ratio (OR): 2.39, 95% Confidence Interval (CI): 1.13, 5.05) and younger age (children <5 years) (OR: 2.09, 95% CI: 1.12, 3.90), and a drinking water source with >1 colony forming unit *E*.*coli* / 100mL (OR: 3.59, 95% CI: 1.46, 8.84) for all *V. cholerae* infections (Table 3). None of the contacts (0 out of 17) residing in households where the index patient consumed doxycycline in the 48 hours prior to healthcare facility admission had a *V. cholerae* infection compared to 28% of contacts (134 out of 474) residing in households where the index patient did not consume doxycycline (p=0.005). None of these contacts consumed doxycycline themselves.

**Table 3.** Logistic Regression Models of *V. cholerae* Infections among 491 Household Contacts of Cholera Patients.

|  | Household<br>Contacts<br>(N=491)<br>% | All Cholera<br>Infections<br>Odds Ratio<br>(95% CI)<sup>1</sup> | All Symptomatic<br>Cholera Infections<br>Odds Ratio<br>(95% CI)<sup>1,2</sup> |
| --- | --- | --- | --- |
| **491 Total Household Contacts of Cholera Patients** |  |  |  |
| **<u>Household Level Characteristics during Surveillance Visits</u>** |  |  |  |
| Index patient antibiotic usage (doxycycline) at baseline (past 48 hours) | 5% | ‡p=0.005 | ‡p=0.400 |
| **Hygiene Indicators** |  |  |  |
| Stored Food Uncovered at All Visits | 26% | **1.822** (1.03, 3.24) | **2.388** (1.13, 5.05) |
| No Soap Present in Cooking Area at All Visits | 14% | 1.406 (0.56, 3.54) | 1.238 (0.44, 3.5) |
| No Soap Present in Toilet Area at All Visits | 56% | 0.809 (0.47, 1.39) | 0.882 (0.42, 1.83) |
| **Water Quality** |  |  |  |
| Stored Household Free Chlorine Concentration <0.2 mg/L at All Visits | 31% | 0.752 (0.43, 1.33) | 0.972 (0.46, 2.07) |
| Source Household Free Chlorine Concentration <0.2 mg/L at All Visits | 47% | 1.292 (0.73, 2.28) | 1.471 (0.68, 3.17) |
| Stored Water with Detectable *V. cholerae* during the Surveillance Period | 11% | 1.172 (0.52, 2.64) | 1.283 (0.57, 2.88) |
| Source Water with Detectable *V. cholerae* during the Surveillance Period | 8% | 0.752 (0.29, 1.92) | 0.892 (0.41, 1.95) |
| Any Water Sample (Stored or Source) with Detectable *V. cholerae* during the Surveillance Period | 16% | 1.389 (0.73, 2.65) | 1.227 (0.62, 2.44) |
| Stored Household *E.coli* Concentration > 1 CFU/ 100 mL during the Surveillance Period | 46% | 0.752 (0.18, 3.21) | 1.998 (0.66, 6.08) |
| Source Household *E.coli* Concentration > 1 CFU/ 100 mL during the Surveillance Period | 23% | **3.594** (1.46, 8.84) | 0.867 (0.39, 1.91) |
| Stored Drinking Water Uncovered at All Visits | 31% | 0.809 (0.46, 1.42) | 0.867 (0.39, 1.91) |
| **Primary Water Source Type<sup>3</sup>** |  |  |  |
| Basic Water Service | 62% | (reference) | (reference) |
| Limited Water Service | 25% | 0.851 (0.42, 1.73) | 0.588 (0.24, 1.44) |
| Unimproved Water Source | 13% | 0.830 (0.34, 2.02) | 0.945 (0.37, 2.42) |
| **Primary Sanitation Option Type<sup>3</sup>** |  |  |  |
| Basic Sanitation Service | 27% | (reference) | (reference) |
| Limited Sanitation Service | 18% | 0.855 (0.39, 1.9) | 0.754 (0.25, 2.27) |
| Unimproved Sanitation | 55% | 0.737 (0.41, 1.34) | 0.735 (0.32, 1.7) |
| **<u>Individual Level Characteristics</u>** |  |  |  |
| Female Household Contacts | 56% | 0.965 (0.71, 1.32) | 0.852 (0.47, 1.53) |
| **Household Contact Patient Age (Years)** |  |  |  |
| 0-5 Years | 20% | 0.955 (0.58, 1.57) | **2.089** (1.12, 3.9) |
| 5-14 Years | 34% | 1.275 (0.83, 1.96) | 1.632 (0.81, 3.27) |
| 14 Years or Greater | 46% | (reference) | (reference) |
| **Blood Group Status** |  |  |  |
| O Blood Group | 44% | 0.881 (0.5, 1.56) | 0.941 (0.44, 2.01) |
| A Blood Group | 31% | (reference) | (reference) |
| B Blood Group | 22% | 0.651 (0.37, 1.15) | 0.619 (0.25, 1.56) |
| AB Blood Group | 3% | 0.333 (0.09, 1.29) | 0.878 (0.13, 5.84) |

## Discussion

This is the first study, to our knowledge, of household transmission of *V. cholerae* in sub-Saharan Africa. Sixty-seven percent of cholera patient households had >1 household contact with a *V. cholerae* infection during the surveillance period. *V. cholerae* was present in both source water and stored household drinking water in cholera patient households. Significant risk factors for symptomatic *V. cholerae* infections were stored food being left uncovered and younger age (children <5 years) with *E*.*coli* in the drinking water source being associated with any type of *V. cholerae* infection. Study findings indicate a high risk of cholera among the household contacts of cholera patients in this urban cholera endemic setting in DRC. These results demonstrate the need for targeted water treatment and hygiene interventions to reduce cholera in transmission hotspots around cholera patients in the DRC.

Twenty-seven percent of household contacts of cholera patient households had a *V. cholerae* infection in our urban setting in the DRC when infection was defined by bacterial culture and serology. This is higher than previous studies in Bangladesh which relied on *V. cholerae* bacterial culture only and found that 20% to 21% of household contacts of cholera patients in an urban setting and 18% in a rural setting had *V. cholerae* infection during the week period after the cholera patient was admitted to a health facility.^8,9,22^ This result is similar to the 19% of contacts with *V. cholerae* infections found in our current study when we relied on bacterial culture data alone during the week after the cholera patient was admitted to a health facility.

Consistent with a previous study we observed higher rates of *V. cholerae* infections when both bacterial culture and serological results were combined. This previous study in Bangladesh found that combining vibriocidal antibody titers with bacterial culture data increased the number of recent *V. cholerae* infections detected by 39% compared to bacterial culture alone (similar to the 42% increase in our current study).^23^

Twelve percent of cholera patient households had a stored water sample with *V. cholerae* and 7% had *V. cholerae* in source water samples at our study site in urban DRC. In our previous study of cholera patient households in urban Bangladesh, 27% of source water samples had *V. cholerae*^22^, more than twice as high as our current finding in the DRC. The percentage of households with stored water with *V. cholerae* was similar, both in our current study in urban DRC at 5% and our previous study in urban Bangladesh at 6%.^22^

Stored food uncovered was a significant risk factor for symptomatic *V. cholerae* infections. We are not aware of another study that has found this association. A meta-analysis identified four studies where consuming a cold meal was associated with an increased risk of *V. cholerae* infections and 3 studies where consuming leftover food was associated with an increased risk of *V. cholerae* infections.^5^ Consistent with our current study these studies suggest that food hygiene plays an important role in *V. cholerae* transmission. Younger age was also significantly associated with an increased risk of a symptomatic *V. cholerae* infection. This finding is consistent with four studies from Uganda, Colombia, India, and Bangladesh.^24-27^ Younger children likely lack the naturally acquired immunity of older individuals that may have already been exposed to a *V. cholerae* infection leading to greater susceptibility to symptomatic *V. cholerae* infections.

No association was observed between *V. cholerae* infections among contacts and *V. cholerae* in stored or source water from cholera patient households. This is in contrast to our previous studies from Bangladesh that observed this association.^9,10^ However, there was an significant association between *E*.*coli* (fecal indicator of water contamination) in the household drinking water source and *V. cholerae* infections among contacts. We are not aware of a previous study that found this association. This finding demonstrates the urgent need for treatment of household drinking water in cholera patient households.

There was no significant association between O blood group status and *V. cholerae* infections among household contacts of cholera patients. However, a significantly higher proportion of index cholera patients had O blood group status compared to their household contacts. This finding suggests that those individuals with O blood group status were more likely to have severe *V. cholerae* infections that required hospitalization. This is consistent with previous studies which have found an association between increased severity of *V. cholerae* infections and O blood group status.^8,23^ Similarly, studies from Bangladesh and Peru finding no association between O blood group status and mild or asymptomatic *V. cholerae* infections.^28,29^

This study has some limitations. First, our study focused only on an urban site in DRC. Future research should be conducted on household transmission of *V. cholerae* in both urban and rural settings in sub-Saharan Africa. Second, we did not perform molecular detection on stool samples which may have increased the number stool samples found to be positive for *V. cholerae* compared to bacterial culture alone.

This study had several strengths. First, the intensive surveillance of cholera patient households which included blood, stool, and water collection at 7 timepoints during the 1-month period after the index cholera patient was admitted to the health facility. Second, we included *V. cholerae* serology to define infection, building on previous studies that relied on bacterial culture data alone from household contacts of cholera patients. Finally, the unannounced spot checks conducted to collect water source and household stored water samples and to assess the presence of soap and the covering status of stored water and food in the home allowed for objective measures of assessing WASH conditions in the household, building on previous studies using self-reported WASH behaviors.

## Conclusion

In this cholera endemic setting in the DRC, the majority of cholera patient households had multiple *V. cholerae* infected household contacts. *V. cholerae* was found in both source water and stored household drinking water in cholera patient households. Furthermore, risk factors for *V. cholerae* infections were stored food left uncovered, younger age (children <5 years), and *E*.*coli* in drinking water sources. Our findings suggest that contamination of drinking water and poor food hygiene practices are potential transmission routes for *V. cholerae* infections in this setting. Therefore, targeted WASH interventions focusing on water treatment and hygiene practices are needed for this high-risk population residing in transmission hotspots for cholera.

## Data Availability

All data produced in the present study are available upon reasonable request to the authors.

## Acknowledgements

We thank the study participants and the following individuals for their support with the implementation of this study: Dr. Placide Welo, Dr. Jean Claude Kulondwa, Dr. Roger Boketsu, Blessing Muderhwa Banywesize, Raissa Boroto, Gisèle Nsimire, Feza Rugusha, Freddy Endeleya, Pacifique Kitumaini, Claude Lunyelunye, Rushago Julienne, Emmanuel Buhendwa, Pascal Kitumaini Bujiriri, Jessy Tumsifu, and Brigitte Munyerenkana. We also thank all staff at the DRC Ministry of Health in South Kivu.

## References

1. Ali M, Nelson AR, Lopez AL, Sack DA. Updated Global Burden of Cholera in Endemic Countries. PLoS neglected tropical diseases 2015; 9(6): e0003832.

2. WHO. Cholera in the WHO African Region: Weekly Regional Cholera Bulletin. 2024. https://www.afro.who.int/health-topics/disease-outbreaks/cholera-who-african-region#:∼:text=As%20of%2031%20July%202024,(4%20375)%20deaths%20reported.

3. Moore SM, Azman AS, Zaitchik BF, et al. El Niño and the shifting geography of cholera in Africa. Proceedings of the National Academy of Sciences of the United States of America 2017; 114(17): 4436–41.

4. Wolfe M, Kaur M, Yates T, Woodin M, Lantagne D. A Systematic Review and Meta-Analysis of the Association between Water, Sanitation, and Hygiene Exposures and Cholera in Case-Control Studies. Am J Trop Med Hyg 2018; 99(2): 534–45.

5. Richterman A, Sainvilien DR, Eberly L, Ivers LC. Individual and Household Risk Factors for Symptomatic Cholera Infection: A Systematic Review and Meta-analysis. J Infect Dis 2018; 218(suppl_3): S154–s64.

6. Ali M, Debes AK, Luquero FJ, et al. Potential for Controlling Cholera Using a Ring Vaccination Strategy: Re-analysis of Data from a Cluster-Randomized Clinical Trial. PLoS Med 2016; 13(9): e1002120.

7. Debes AK, Ali M, Azman AS, Yunus M, Sack DA. Cholera cases cluster in time and space in Matlab, Bangladesh: implications for targeted preventive interventions. Int J Epidemiol 2016.

8. Weil AA, Khan AI, Chowdhury F, et al. Clinical outcomes in household contacts of patients with cholera in Bangladesh. Clinical infectious diseases 2009; 49(10): 1473–9.

9. George CM, Hasan K, Monira S, et al. A prospective cohort study comparing household contact and water Vibrio cholerae isolates in households of cholera patients in rural Bangladesh. PLoS Negl Trop Dis 2018; 12(7): e0006641.

10. Burrowes V, Perin J, Monira S, et al. Risk Factors for Household Transmission of Vibrio cholerae in Dhaka, Bangladesh (CHoBI7 Trial). The American journal of tropical medicine and hygiene 2017; 96(6): 1382–7.

11. George CM, Namunesha A, Felicien W, et al. Evaluation of a rapid diagnostic test for detection of Vibrio cholerae O1 in the Democratic Republic of the Congo: Preventative intervention for cholera for 7 days (PICHA7 program). Tropical medicine & international health : TM & IH 2024.

12. Halder AK, Tronchet C, Akhter S, Bhuiya A, Johnston R, Luby SP. Observed hand cleanliness and other measures of handwashing behavior in rural Bangladesh. BMC public health 2010; 10: 545.

13. Organization WH. Guidelines for drinking-water quality: incorporating the first and second addenda: World Health Organization; 2022.

14. WHO G. Guidelines for drinking-water quality. World Health Organization 2011.

15. WHO/UNICEF. The JMP ladder for sanitation. 2020. https://washdata.org/monitoring/sanitation.

16. JMP/UNICEF. The JMP service ladder for drinking water. 2022.

17. Islam MS, Siddika A, Khan M, et al. Microbiological analysis of tube-well water in a rural area of Bangladesh. Applied and environmental microbiology 2001; 67(7): 3328–30.

18. Monsur K. Bacteriological diagnosis of cholera under field conditions. Bulletin of the World Health Organization 1963; 28(3): 387.

19. Organization WH. Manual for the laboratory identification and antimicrobial susceptibility testing of bacterial pathogens of public health importance in the developing world: Haemophilus influenzae, Neisseria meningitidis, Streptococcus pneumoniae, Neisseria gonorrhoea, Salmonella serotype Typhi, Shigella, and Vibrio cholerae: World Health Organization, 2003.

20. Daniels G, Reid MEJT. Blood groups: the past 50 years. 2010; 50(2): 281–9.

21. Leung DT, Uddin T, Xu P, et al. Immune responses to the O-specific polysaccharide antigen in children who received a killed oral cholera vaccine compared to responses following natural cholera infection in Bangladesh. Clin Vaccine Immunol 2013; 20(6): 780–8.

22. George CM, Monira S, Sack DA, et al. Randomized Controlled Trial of Hospital-Based Hygiene and Water Treatment Intervention (CHoBI7) to Reduce Cholera. Emerg Infect Dis 2016; 22(2): 233–41.

23. Harris JB, Khan AI, LaRocque RC, et al. Blood group, immunity, and risk of infection with Vibrio cholerae in an area of endemicity. Infection and immunity 2005; 73(11): 7422–7.

24. Cárdenas V, Saad C, Varona M, Linero M. Waterborne cholera in Riohacha, Colombia, 1992. Bulletin of the Pan American Health Organization 1993; 27(4): 313–30.

25. Cummings MJ, Wamala JF, Eyura M, et al. A cholera outbreak among semi-nomadic pastoralists in northeastern Uganda: epidemiology and interventions. Epidemiology and infection 2012; 140(8): 1376–85.

26. Saha A, Hayen A, Ali M, et al. Socioeconomic risk factors for cholera in different transmission settings: An analysis of the data of a cluster randomized trial in Bangladesh. Vaccine 2017; 35(37): 5043–9.

27. Sur D, Deen JL, Manna B, et al. The burden of cholera in the slums of Kolkata, India: data from a prospective, community based study. Archives of disease in childhood 2005; 90(11): 1175–81.

28. Glass RI, Holmgren J, Haley CE, et al. Predisposition for cholera of individuals with O blood group. Possible evolutionary significance. American journal of epidemiology 1985; 121(6): 791–6.

29. Swerdlow DL, Mintz ED, Rodriguez M, et al. Severe life-threatening cholera associated with blood group O in Peru: implications for the Latin American epidemic. The Journal of infectious diseases 1994; 170(2): 468–72.

