## Supplemental File 1 for "Epidemiology of *Vibrio Cholerae* Infections in the Households of Cholera Patients in the Democratic Republic of the Congo: PICHA7 Prospective Cohort Study"

**Supplemental File 1.** Serum Antibody Analysis via ELISA

Serum was separated from blood and frozen at -80°C. Serum samples were shipped to the United States and analyzed for IgG and IgA antibodies to *V. cholerae* O1 Ogawa and Inaba O-specific polysaccharide (OSP), and IgG to cholera toxin B subunit (CTB) using enzyme-linked immunosorbent assay (ELISA) adapted from previously published methods (1). Briefly**,** using the Tecan Freedom EVO 96-well liquid handling system, Nunc Maxisorp 384 well plates (Invitrogen) were coated with 1.0 μg/ml OSP:BSA (gift from Dr. Edward Ryan, Massachusetts General Hospital, Boston, MA) in carbonate buffer and incubated overnight at 4°C. For CTB ELISAs, plates were first coated with Monosialoganglioside GM1 (Sigma-Aldrich) (1.0 μg/ml) in carbonate buffer overnight, subsequently blocked and incubated overnight with CTB (Sigma-Aldrich) (2.5 μg/ml). All plates were then blocked and serum (diluted 1:10 in BioStab antibody stabilizer (Sigma-Aldrich), further diluted 1:10 in 0.1% BSA-PBS-tween (0.05%)) was added to the plate to incubate for 2 hours at room temperature (RT). Plates were incubated with 1:1000 dilution of anti-human IgG or IgA (Invitrogen) HRP conjugate for 2 hours at RT. Plates were developed by using the substrate 3,3′,5,5′-tetramethylbenzidine (Sigma) and were read kinetically at 405nm for 20 minutes at 2-minute interval using a BioTek Synergy plate reader (Agilent). ELISA units were normalized to a monoclonal antibody standard (gift from Dr. Richelle Charles, Massachusetts General Hospital, Boston, MA).

1.	Leung DT, Uddin T, Xu P, Aktar A, Johnson RA, Rahman MA, et al. Immune responses to the O-specific polysaccharide antigen in children who received a killed oral cholera vaccine compared to responses following natural cholera infection in Bangladesh. Clin Vaccine Immunol. 2013;20(6):780-8.
